# PROTOCOL – Are investigators’ access to trial data and rights to publish restricted and are trial participants informed about this? A comparison of trial protocols and informed consent materials

**DOI:** 10.1101/2020.08.13.20174177

**Authors:** Asger Sand Paludan-Müller, Karsten Juhl Jørgensen, Peter C. Gøtzsche

**Author notes:** Corresponding author Asger Sand Paludan-Müller, Nordic Cochrane Centre, Rigshospitalet, Dept. 7811, Blegdamsvej 9, 2100 Copenhagen Ø.

## Abstract

This is a protocol for the project entitled: “Are investigators’ access to trial data and rights to publish restricted and are trial participants informed about this? A comparison of trial protocols and informed consent materials”

## Background

Cooperation between pharmaceutical companies and academic investigators is common for randomised clinical trials (RCTs) [1,2]. While this construct has advantages it is essentially a business transaction and conflicts of interest are common. There is convincing empirical evidence of selective reporting of results [1,3], and industry trials are less likely to be published than non-industry trials [4,5].

Previous studies have examined constraints on publication rights in industry-initiated trials. In 2006, a study found that out of 44 trials approved by an ethics committee in Denmark in 1994-1995, 40 trials (91%) described constraints on publication in the trial protocol and the same was true for 41 out of 44 trials (93%) approved in 2004 [6]. In 2016, a study examined whether there were constraints on publication in 647 protocols approved by ethics committees in Switzerland and Germany between 2000 and 2003. 456 trial protocols mentioned publication agreements and in 393 of those (86.2%) the industry partner had the right to either disapprove or at least review publications [7],

Both studies used relatively old samples, and to our knowledge no study has examined publication constraints in a recent sample of RCTs approved by ethics committees. Additionally, none of the previous studies have compared information on publication restraints available to ethics committees with the information provided to research participants. As altruism is generally believed to be an important reason for participating in clinical trials [8,9], we consider it important that patients are informed of potential publication constraints.

Another potentially problematic issue in clinical trials is early stopping. A 2010 review found that trials that stopped early for benefit exaggerated the effect by 29% compared to trials of the same intervention that had not stopped early [10]. In the 2006 study, the industry sponsor had access to accumulating data in 16 out of 44 trials (36%) and the sponsor could stop the trial at any time for any reason in an additional 16 trials (36%).

We wish to examine to what degree access to data and the right to publish is restricted, whether this is communicated to patients, and whether the industry partner has the opportunity to accumulate data and stop the trial early, using a sample of relatively recent RCTs approved by ethics committees in Denmark.

### Research questions

- Are investigators’ access to data and right to publish constrained in clinical trials with industry involvement approved by ethics committees in Denmark?
- Are any such constraints communicated to research participants?
- Does the industry partner have the opportunity to accumulate data during the trial and stop the trial early?

## Methods

### Retrieval of trial protocols

This project will be based on a sample of protocols and other documents submitted to research ethics committees in Denmark for 67 trials. The sample has been used for other related projects, and the process of obtaining access to the documents through a Freedon of Information enquiry has been described in details elsewhere [11,12],

### Data extraction

#### Trial characteristics

One observer will extract trial characteristics from the protocols, and if necessary other documents, for all included trials. We will extract the following information:

- Title
- Medical condition studied
- Medical speciality
- Experimental intervention and comparator(s) used, including dosing schedules
- Number of arms
- Single-site or multi-centre study
- Planned sample size
- Funding source(s)
- Trial duration
- Primary outcome(s)
- Trial phase (not relevant for trials studying procedures or non-medicinal products).

The trial characteristics will be entered into an Microsoft Excel spreadsheet [16].

#### Information on roles, responsibilities and rights of investigators, sponsors, and funders

One observer will extract all information on the roles, responsibilities, and rights of investigators, sponsors, and funders, including information on access to data and rights to publish. We will also extract information about the sponsors access to accumulating data during the study (e.g. through interim analyses) and early stopping rules. This information will be extracted from protocols and other relevant documents submitted to the ethics committees, such as publication agreements and contracts.

We will also extract any information on ownership of data and rights to publish provided to research participants in Informed Consent Documents (ICDs)

### Analysis

Using the information in the dedicated spreadsheet, one observer will assess the following domains for all included trials.

1. *Are the roles and responsibilities of the trial funders and sponsors described ?* We will assess whether the role and responsibility of trial funders and sponsored are reported in detail in the protocols and, if yes, note what these roles and responsibilities are.
2. *Does the industry partner own the data ?* We will assess statements about who owns the data, and whether there are any restrictions to investigators’ access to data.
3. *Were the investigators’ rights to publish restricted?* We will assess whether the investigators’ rights to publish are stated to be restricted and, if yes, what the nature of such restrictions is.
4. *Did the industry partner have access to accumulated data during the study?* We will assess if any procedures for the sponsor to access accumulated data during the study are described (e.g. interim analyses or participation in data and safety monitoring committees) and, if yes, the nature of the procedures.
5. *Were early stopping rules described?* We will assess if any rules for early stopping of trials is described and, if yes, under what circumstances the trials can be stopped.
6. *Was information about ownership of data, stopping rules and restrictions on investigators’ right to publish provided to trial participants in ICDs?*

If we identify any restrictions on access to data or rights to publish for a given trial, we will examine whether research participants are made aware of them.

### Reporting and dissemination

This protocol will be made available from the medRxiv preprint server. The results will be published in a peer-reviewed medical journal.

## Data Availability

Due to confidentiality issues, the data from this project will not be made available

## Contributions

PCG conceived the idea and wrote the study proposal. KJ and ASP contributed to the design of the study. ASP wrote the first draft of this protocol, and all authors revised the protocol and approved the final version.

